# Physical Profile and Epidemiological Analysis of Injuries in a College Athletics Team: A Retrospective Analysis

**DOI:** 10.1101/2021.11.02.21265630

**Authors:** Pedro Rocha Tenorio, Jean Kleber de Oliveira Santos, Mariana Emanuela Higa de Melo, Thaoan Bruno Mariano

## Abstract

**Introduction:** Athletics is a sport based on natural patterns and activities. As a sport, presents an array of benefices such as the improvement of physical condition and personal interaction, however, is not free of risks such as injuries related to training and competition.

**Objectives:** Analyze the physical profile and associated sports injuries in an athletics college team.

**Methods:** Subjects enrolled in a medicine college athletics team from Jan to Oct of 2021 of both genders had their characteristics collected and answered to a survey modified from the “NCAA Injury Surveillance System”.

**Results:** 31 subjects answer the survey. 65% of the subjects present at least 1 injury, 73.9% of injuries were classified as severe, 0.27 injuries were reported per 1000 hours of exposure. Injured athletes had a practice time 2-fold greater than uninjured athletes. Quadriceps and shin injuries represent 52.17% of the injuries.

**Conclusion:** College athletics seems to present a high risk of severe sports injuries in the lower body, and the greater risk factor seems to be the practice time.

## INTRODUCTION

The practice of physical activities has a number of recognized health benefits, such as fitness improvement, disease prevention, reduction of drug abuse, personal and social development^1^. However, there are negative effects related to sports, especially in subjects that aim high performance, such as eating disorders^1^ and sports injuries^2^.

The injury rate of athletes is greater than in the general population, varying according to the modality, training volume, specialization, and other factors^2,3,4^. The time away from the sport due to injuries can vary depending on the site and severity of the injury, as well as the type of treatment adopted, ranging from a few days^5^ to months in surgical cases^6^.

Although college athletics in Brazil are not expressive as in other countries, like the US where the practices are related to professional careers and scholarships^7^, there are sports events organized by the Brazilian Confederation of College Sports (CBDU) in which athletes compete in a variety of athletics modalites^8^.

Among Brazilian research the scientific production related to athletics are scarce^6,7^. The medicine college from the University of the West of São Paulo (UNOESTE), Brazil, has an athletics team, however, there is no description of these individuals, therefor the present study accomplished an epidemiological survey of the physical characteristics and injuries presented by the athletes, in addition to identify risk factors associated with the injuries.

## METHODS

### Ethical Considerations

This study followed all ethical standards of experimentation and was approved by the Research Ethics Committee and registered with the Research, Development and Innovation Coordination of the University of the West of São Paulo under number 6991.

### Patients and Public Involvement

Patients or the public were not involved in the design, conduct, reporting, or dissemination plans of this research.

### Design

A retrospective analysis of the UNOESTE athletics team was performed. Subjects were allocated individually in a room with a researcher, data of interest were collected and a survey was delivered to the participants. To avoid biases in completing the survey, participants were previously instructed and the researchers were available to clarify any doubts while answering the survey.

### Recruitment and Data Collection

All individuals enrolled from January to October 2021 in the athletics team, from both genders, were invited to participate in the study. Participants’ height and weight were collected using a digital scale and a measuring tape. A modified version of the Injury Surveillance System (ISS)^8^ was applied, containing questions about the practice time, modality practiced, site, date and mechanism of injury, physical and clinical therapeutic diagnosis, treatment adopted, and time away after injury.

### Athlete Exposures

To determine the injury rate per 1000 hours of exposure (AE), the weekly training time and sport practice time plus the time in competitive events was determinate. The total number of injuries was then divided by the total exposure time and multiplied by 1000, resulting in the AE.

### Data Analysis

Shapiro-Wilki test was performed to check for normality. Age, height, weight, body mass index (BMI), weekly training and total practice time were compared according to gender and presence of injured using independent sample Student t-test. The Risk Ratio (RR) with 95% confidence interval (95%IC) and χ^2^ test was applied to assess the relationship between gender and the occurrence of injuries. A significance level of 5% was assumed. The software used was SPSS Statistics (V.20, IBM, Armonk, New York, USA).

## RESULTS

In total 32 participants are invited and 31 answer the survey. The study flowchart is shown in Figure 1.

### Subject Characterization

Population characteristics are presented in Table 1. Due to condition standardization issues, the body fat percentage data were discarded. No other data was lost.

**Table 1.**
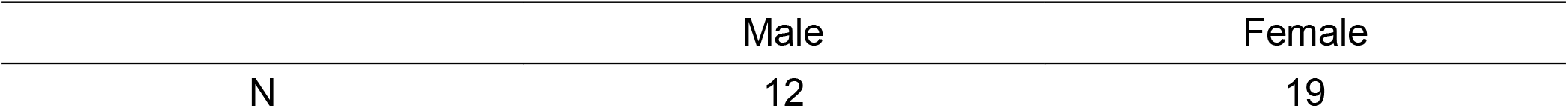
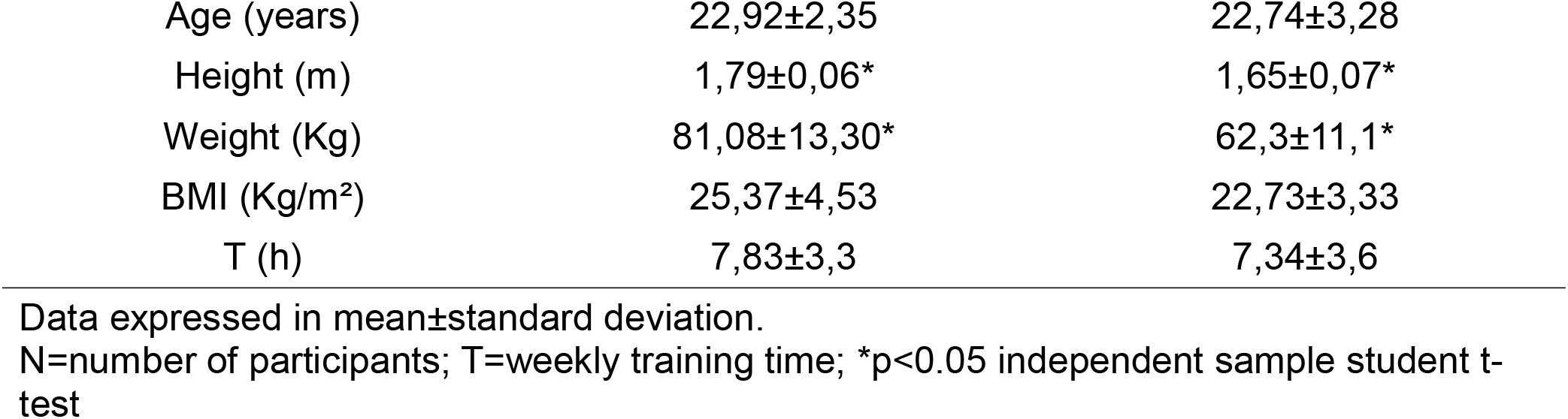
Population Characterization

The number of modalities practiced per athlete ranged from 1 to 6, and their distribution is presented in Table 2.

**Table 2.** Number of Participants per Modality

| Modality | Participants<br>(Male/Female) |
| --- | --- |
| 100 m | 7/12 |
| 400 m | 4/5 |
| 1500 m | 2/4 |
| 5000 m | 2/0 |
| 4x100 m relay | 5/4 |
| 4x400 m relay | 4/0 |
| Shot Put | 2/5 |
| Disc Throw | 1/2 |
| Javelin Throw | 3/0 |
| Long Jump | 1/2 |
| High Jump | 2/1 |
| Triple Jump | 2/0 |

Only 6 participants had competition history, 3 competed once, 2 competed twice, and 1 competed 5 times. All competing at the regional or state level. 4 of the participants with competition history had injuries, with the 2 uninjured individuals having competed only 1 time.

### Injury Assessment

A total of 23 injuries were found, with 17 athletes sustaining 1 while 3 athletes sustaining 2 injuries. 52.6% of women and 83.3% of men had at least 1 injury. No association was found between gender and injury (RR, 1.58; 95%IC, 0.96-2.59; p=0.06). The overall exposure time was 82627.62 hours, resulting in 0.28 AE, with no difference in a gender sub-analysis.

The site of injuries are presented in Table 3.

**Table 3.** Incidence of injuries by site

| Injury Site | N (%) |
| --- | --- |
| Shin | 7 (30.43) |
| Quadriceps | 5 (21.74) |
| Hamstrings | 3 (13.04) |
| Knee | 3 (13.04) |
| Patella | 2 (8.69) |
| Ankle | 2 (8.69) |
| Shoulder Blade | 1 (4.35) |

N=number of injury

All recorded injuries occurred during training: 2 during warm-up, 9 during the first half, 7 during the second half, and 5 during the cool-down. 20 were new injuries, 2 were recurrent injuries from the same modality and 1 was due to a complication of a previous injury from another modality. 2 of the injuries led to the participants’ permanent removal, 3 injuries didn’t result in time away and the average time away for the other injuries was 53.18 days, varying from 7 days to 6 months. The types of injuries can be seen in Figure 2.

Only 13 (56.3%) injuries were clinically diagnosed, and imaging exams were performed in 8 of them. 2 injuries were treated with non-steroidal anti-inflammatory medication for 5 and 7 days, and 2 were treated with prescribed dietary supplementation for 3 and 5 months. None of the injuries underwent surgical intervention. 11 (47.8%) injuries were treated with physical therapy, of which 9 of them had a mean treatment time of 1.25±0.47 months and another 2 had a discrepant time of 6 to 16 months.

### Risk Factor

The characterization of injured and uninjured individuals is shown in Table 4.

**Table 4.** Characteristics of injured and uninjured participants

|  | Injured (20) | Uninjured (11) |
| --- | --- | --- |
| Age (years) | 22,45±2,19 | 23,45±3,3,96 |
| Height (m) | 1,72±0,1 | 1,68±0,08 |
| Weight (Kg) | 67,58±11,31 | 73,18±20,27 |
| BMI (Kg/m<sup>2</sup>) | 22,65±2,76 | 25,74±5,15 |
| Weekly Training (h) | 7,32±3,34 | 7,91±3,75 |
| Total Practice Time (months) | 15,1±10,66* | 8±5,19* |

Data expressed in mean ± standard deviation.
\*p=0.048 independent sample student t-test.

## DISCUSSION

### Incidence and severity of injuries

This study found a high prevalence of injuries in athletes from a college center, with an unexpected high incidence of injuries classified as severe, defined as more than 21 days of time away^9^.

In a study by Lambert *et al*. conduced retrospectively with 743 professional athletics athletes that answered a questionnaire formulated by the German Olympic Federation, where participants self-reported their injuries occurrence during the 2012-2016 Olympic cycle, found that 64 % of participants had injuries, a similar level that reported in this study^10^.

The injury rate was also similar to the reported in a recent systematic review conducted by Francis *et al*., which summarized data from 36 studies, totaling 18,195 runners^11^. As running was the main modality practiced by the present group, this may explain the higher incidence of injuries compared to other studies in athletics with greater variety of modalities^3^.

The incidence of serious injuries was much higher than that reported by Kay *et al*. in the NCAA Injury Surveillance Program (NCAA ISP), a prospective study that analyzed data across the affiliated centers. In the sum of the subgroups of athletics, the authors founded during the academic period of 2009-10 to 2014-15, a total of 220 severe injuries out of 1881, about 11.7%, compared to 73.9% reported in this study^9^.These data may reflect the type of injuries reported, such as ruptures, fractures and cartilage damage. The prolonged time away can have a deleterious effect on the participants’ lives as a whole, preventing athletes from participating not only in sports activities, but also in social and academic events, compromising their physical and mental health.

Although the total incidence of injury in athletes was higher than expected, the AE was low. In a Swedish work carried out by Zachrisson *et al*. which followed during 1 season, 59 athletes of both genders with an average age of 21.6 years, found an incidence of 1.81 AE^12^, 6.7 times higher incidence than that reported in this study.

Boltz *et al*. reported similar values in another work derived from the NCAA ISP, which assessed 1081 male athletes during 455,609 exposure hours during the period of 2014-15 to 2018-19, which found a rate of 2.37 AE^13^. This difference may be due to the amateur level and the small number of competitions held by the participants, which theoretically would reflect a lower training intensity and a low exposure of events under stress conditions, favoring a lower rate of AE, data corroborated by the history of competition and injuries presented in this study, which indicates a greater chance of injury with a greater number of competitions.

Injury recurrence was small in contrast to that reported in a cross-sectional analysis of college-level athletes conducted by Lemoyne *et al*. where 82 participants answered a questionnaire in which they found an incidence of more them 2 injuries per injured athlete (IIA)^14^. Compared to the study by Lambert *et al*., which found an incidence of 1.32 IIA^10^, this study presents similar findings with an incidence of 1.15 IIA. It is believed that the low recurrence of injuries was due to the long time away reported by athletes after an injury, which can favor a complete recovery, avoiding recurrence or complications of the injuries. These findings demonstrate the need for careful reasoning when deciding to reintegrate a participant after an injury, considering the appropriate balance between the negative and positive effects of time away.

### Injury Site

The injuries per body segment were similar to that found by Lamberte *et al*., who reported 83% of injuries in the lower limbs^10^, compared to 95.6% reported in the present study. The anatomical sites of injuries were also similar. The findings are consistent with the study conducted by Francis *et al*.^11^ when compared to lesions on the lower limbs. These data indicate that the lower body of athletics athletes, especially thighs, knees and ankles, are the region with the greatest effort demand and susceptibility to injuries.

### Risk Factors

The only significant factor was the total practice time, being almost 2-fold greater in injured athletes, corroborating the literature, as longer practice time corresponds to a greater number of exposure hours^15^, and a greater accumulated training load, recognized risk factors for the development of injuries^2,16^.

### Starting Problem Solving

The high rate of injuries and the agreement with the literature regarding the critical areas affected demonstrate the need to implement a targeted prevention and rehabilitation program, a measure of vital importance for the health and longevity of athletes.

Approaches such as the one described by Macdonald *et al*.^17^, providing a process of classification, rationalization, joint decision-making as well as prescription and progression of treatment, proved effective in subsequent work of Pollock *et al*.^18^, being able to reduce the time away and the recurrence of injuries, are excellent starting points.

### Limitations

The limitations present in the study are due to its retrospective nature, which may lead to under-reporting of minor injuries that may have been forgotten, as the accuracy of the reported data. The inaccuracy in the report of a second sport was evident, individuals reported weight training for intermittent periods but were unable to determine the period, as well as the low sample number, which may have interfered in the identification of risk factors. Prospective studies in order to confirm the findings and validate the implementation of targeted preventive measures are needed.

## CONCLUSION

The athletics team analyzed showed a low injury per exposure time with a high prevalence of injuries, especially in the lower limbs, of a severe nature, and its main risk factor seems to be the training practice time.

## Data Availability

All data produced in the present study are available upon reasonable request to the authors

